# Communicative and critical health literacy and eating behaviors in Japanese adults: the modifying role of body image distortion

**DOI:** 10.1101/2025.08.01.25332557

**Authors:** Noriaki Kurita, Takako Maeshibu, Sayaka Shimizu, Tetsuro Aita, Takafumi Wakita, Hiroe Kikuchi

## Abstract

**Background:** Disordered eating behaviors, emotional eating (EE), uncontrolled eating (UE), and cognitive restraint (CR) contribute to weight dysregulation and remain public health concerns in Japan. Body image distortion (BID), or the misperception of one’s body size, has been linked to both being underweight and overweight. While health literacy (HL) and BID are individually associated with eating behaviors, the influence of higher-order HL domains—communicative and critical literacy—and their interactions with BID remain unclear. This study examined how HL and BID jointly influence multidimensional eating behaviors.

**Methods:** In this cross-sectional study of Japanese adults, HL was measured using the 14-item Health Literacy Scale. BID was defined as the discrepancy between the perceived (via the Figure Rating Scale) and actual body size (via BMI)and categorized as underestimation, no distortion, or overestimation. Eating behaviors (UE, EE, and CR) were assessed using the Japanese version of the 18-item Three-Factor Eating Questionnaire-R18V2. Multinomial logistic regression was used to examine the association between HL and BID, and general linear models were used to test whether BID moderated the effects of higher-order HL domains on eating behavior.

**Results:** Among the participants, 13.0% were underweight, 60.2% had a normal weight, 18.7% were overweight, and 8.0% were obese. BID was categorized as overestimation (36.7%), non-distortion (53.6%), or underestimation (9.7%). HL scores were not significantly associated with overestimation or underestimation; however, a higher BMI was inversely related to overestimation. Higher functional HL was associated with lower EE, UE, and CR across all BID subgroups. However, associations for communicative HL varied by BID (interaction P = 0.001 for EE, 0.057 for CR); it was positively associated with EE and CR in the underestimated group, but inversely associated with EE in the overestimated group. Critical HL was positively associated with CR in the overestimation group (interaction, P = 0.018).

**Conclusion:** Fostering functional HL may support healthier eating behaviors, regardless of BID. Communicative HL may be beneficial for individuals with overestimation-type BID, but potentially counterproductive for those with underestimation. Critical HL appears to encourage more restrictive eating, specifically among individuals with overestimated HL.

## Background

Disordered eating behavior is a key factor that contributes to difficulties in maintaining a healthy weight. Emotional eating (EE)—overeating triggered by negative emotions—is a known risk factor for overweight and obesity and represents a potential target for intervention [1,2] Uncontrolled eating (UE), defined as general difficulties in regulating eating, has been linked to increased body mass index (BMI) and disordered eating patterns, such as binge eating. [3]. Cognitive restraint (CR), the conscious restriction of food intake to control weight, may play an adaptive role in obesity management [4]. However, it has also been biologically associated with underweight status in women and [5], paradoxically, with increased BMI when excessive. [4,6]. Regulating these eating behaviors is particularly important in light of widespread public health concerns regarding underweight and obesity among Japanese adults. According to the 2023 National Health and Nutrition Survey, 24.4% of women aged 20–29 years are underweight, while obesity affected 5.6% of women and 5.9% of men aged 20–69 years [7]. Despite the potential influence of individual cognitive factors, such as health literacy (HL) and body image (BI), on eating behavior, particularly through their interactive effects, few studies have examined these associations in adult populations.

HL—the ability to obtain, understand, and apply health-related information—has been linked to eating behaviors in populations such as Turkish adults and Iranian nursing students. [8,9] Beyond basic functional literacy, HL includes higher-order domains such as communicative and critical literacy [10]. In Japanese adults, lower HL levels, including communicative and critical domains, have been associated with a higher likelihood of unhealthy behaviors such as regular drinking and physical inactivity [11]. However, the relationship between these higher-order domains and multidimensional eating behavior remains poorly understood.

Body image distortion (BID), perceiving oneself as thinner or heavier than one’s actual body size, can contribute to both being overweight and underweight. BI underestimation has been linked to higher rates of obesity [12], whereas overestimation is associated with lower BMI in young to middle-aged women in Japan and increased disordered eating symptoms among adolescents in Slovakia [13, 14]. Moreover, higher HL has been correlated with greater concern for or dissatisfaction with BI [15], suggesting that the combination of higher-order HL and BID may influence multidimensional eating behaviors in different ways.

This study examined the interrelationships between higher-order HL, BID, and multidimensional eating behaviors among Japanese adults across a broad age range. A better understanding of these complex associations may help inform tailored interventions targeting specific HL domains based on individual BID patterns to promote healthier eating behaviors.

## Method

### Aim, Design, and Setting

This study aimed to investigate the joint association between HL and BID and multidimensional eating behaviors in a diverse sample of adult Japanese men and women. A cross-sectional online survey was conducted with the assistance of a web-based research company (Cross Marketing Inc., Tokyo, Japan) and was approved by the institutional review board of our university (approval number: ippan2022-210). The methodological framework is described in our previous work [16]. Participants (N = 1,500) were recruited using stratified sampling based on sex (1:1), age group (<65 vs. ≥65), and obesity-related characteristics (treatment history, weight concerns, or neither). The detailed stratification procedures are provided in Additional File 1. Participants completed the online questionnaire between January 26 and 31, 2023, and received incentive points redeemable for cash, gift certificates, or mileage.

### Participant Screening and Data Quality

To minimize careless or invalid responses, five exclusion criteria were applied: inappropriate or inconsistent entries for (1) age or (2) sex, extreme values for (3) height or (4) weight, and (5) survey completion time < five minutes. No missing data were present, as all items were mandatory. Respondents were asked to report their age and sex at both the beginning and end of the survey, and inconsistent responses led to their exclusion. Extreme height or weight values were excluded, and no additional BMI-based cutoffs were applied. All the remaining responses were retained for analysis.

### Exposure: Multidimensional Health Literacy

Health literacy was assessed using the 14-item Health Literacy Scale (HLS-14), a self-administered instrument developed for the general Japanese population [17]. The scale comprises three subscales: functional HL (five items), communicative HL (five items), and critical HL (four items). Each item was rated on a 5-point Likert scale ranging from 1 (“strongly disagree”) to 5 (“strongly agree”), with the five functional items reverse-scored. The subscale scores were calculated by summing the item responses within each domain, with higher scores indicating higher health literacy. The reliability and validity of the HLS-14 have been previously established in a Japanese population [17].

### Body Image Distortion (BID)

BID refers to the discrepancy between one’s perceived and actual body size, which is either underestimated or overestimated. Perceived body size was assessed using the Figure Rating Scale, which presents nine sex-specific silhouettes ranging from very thin (1) to very large (9) [18, 19] (see Additional File 2). The participants selected the figure that best represented their current body shape. Following a previous study [20], responses were categorized as underweight (1–2), normal weight (3–4), overweight (5–7), or obese (8–9). Actual body size was calculated using BMI, defined as weight (kg) divided by height squared (m²), and categorized as underweight (<18.5), normal weight (18.5–24.9), overweight (25.0–29.9), or obese (≥30.0).

BID was determined by comparing the FRS-based body size with the BMI-based classification.

Underestimate: perceived body size < actual BMI category

No distortion: perceived body size = actual BMI category

Overestimate: perceived body size > actual BMI category

### Outcome: Eating Behavior (Three-Factor Eating Questionnaire-R18V2)

Eating behavior was assessed using the Japanese version of the 18-item Three-Factor Eating Questionnaire-R18 version 2 (TFEQ-R18V2), originally developed by Cappelleri and Karlsson [21]. With permission from the original developer (Karlsson), two physicians with experience in scale development translated the questionnaire into Japanese. The translation was then back-translated into English by two bilingual translators (one American and one Canadian), followed by minor revisions based on comparisons with the original. The final version was reviewed and approved by the original author ( Additional File 3).

Participants were instructed to read each of the 18 items carefully and select the option that best applied to them, using a 4-point Likert scale. Each item is scored on a scale of 1 to 4. Consistent with the original scale, items 1–16 were reverse coded before analysis [21]. Domain scores were calculated as transformed scores ranging from 0 to 100 by subtracting the lowest possible raw score from the total score, dividing it by the possible score range, and multiplying it by 100. Higher scores indicate greater levels of cognitive restraint (CR; three items), uncontrolled eating (UE, nine items), and emotional eating (EE, six items).

### Other Survey Variables

A detailed explanation of item selection is provided in Additional File 4.

The Japanese version of the Dutch Eating Behavior Questionnaire (DEBQ), a 33-item scale measuring emotional, external, and restrained eating, was used to assess eating behaviors [22, 23]. Participants rated each item on a 5-point Likert scale ranging from 1 (never) to 5 (very often), and subscale scores were calculated by averaging the corresponding items [23]. Higher scores indicate stronger tendencies toward the respective eating behaviors. The alpha coefficients for each domain were as follows: emotional eating, 0.95; external eating, 0.73; restrained eating, 0.87 [22].

Demographic and clinical covariates included age, sex, educational attainment, household income, marital status, exercise habits, and history of psychiatric conditions (e.g., eating disorders, depression, and other psychiatric disorders).

### Statistical analysis

Psychometric evaluations were conducted using R (version 4.1.2), employing the psych (v2.2.3) and lavaan (v0.6-11) packages. All the remaining statistical analyses were performed using Stata/SE version 17 (StataCorp, College Station, TX, USA). Participant characteristics were described using means and standard deviations (SDs) or median and interquartile range (IQR) for continuous variables, and frequencies and percentages for categorical variables.

To assess the structure of the TFEQ-R18V2, confirmatory factor analyses were conducted based on a three-factor model using appropriately recoded item scores (Items 1–16). Model fit was evaluated using the comparative fit index (CFI ≥ 0.90) and the root mean square error of approximation (RMSEA ≤ 0.08) [24]. Factor loadings were considered acceptable if ≥ 0.30 [25]. We further explored item-level distributions to detect potential floor or ceiling effects, defined as more than 50% of the responses clustered at either extreme [21]. Internal consistency within each domain was estimated using Cronbach’s α and McDonald’s ω coefficients [26]. To examine the construct validity, correlations between the TFEQ-R18V2 and the corresponding domains of the DEBQ were calculated. Criterion validity was assessed by correlating TFEQ-R18V2 scores with BMI using Spearman’s correlation coefficients, as appropriate. The theoretical basis for the expected correlations is outlined in Additional File 5.

We applied general linear models adjusted for the above-mentioned covariates to examine the associations among multidimensional HL, BID, and eating behavior. To explore potential effect modification by BID, interaction terms were added separately for each HL domain—functional, communicative, and critical HL. Two interaction terms were included in each domain-specific model, and global interaction effects were assessed using the Wald test. These analyses were conducted independently for each of the three domains of the TFEQ-R18V2: EE, UE, and CR. As all survey items were mandatory in the online questionnaire, there were no missing data. Statistical significance was defined as P < 0.05 for all analyses, except for global interactions evaluated by the Wald test, where P < 0.10 was considered indicative of significance.

## Results

A total of 2,155 individuals participated in this study. After excluding 514 respondents with inattentive or implausible responses, 1,641 were included in the primary analysis (Figure 1).

**Figure 1.**
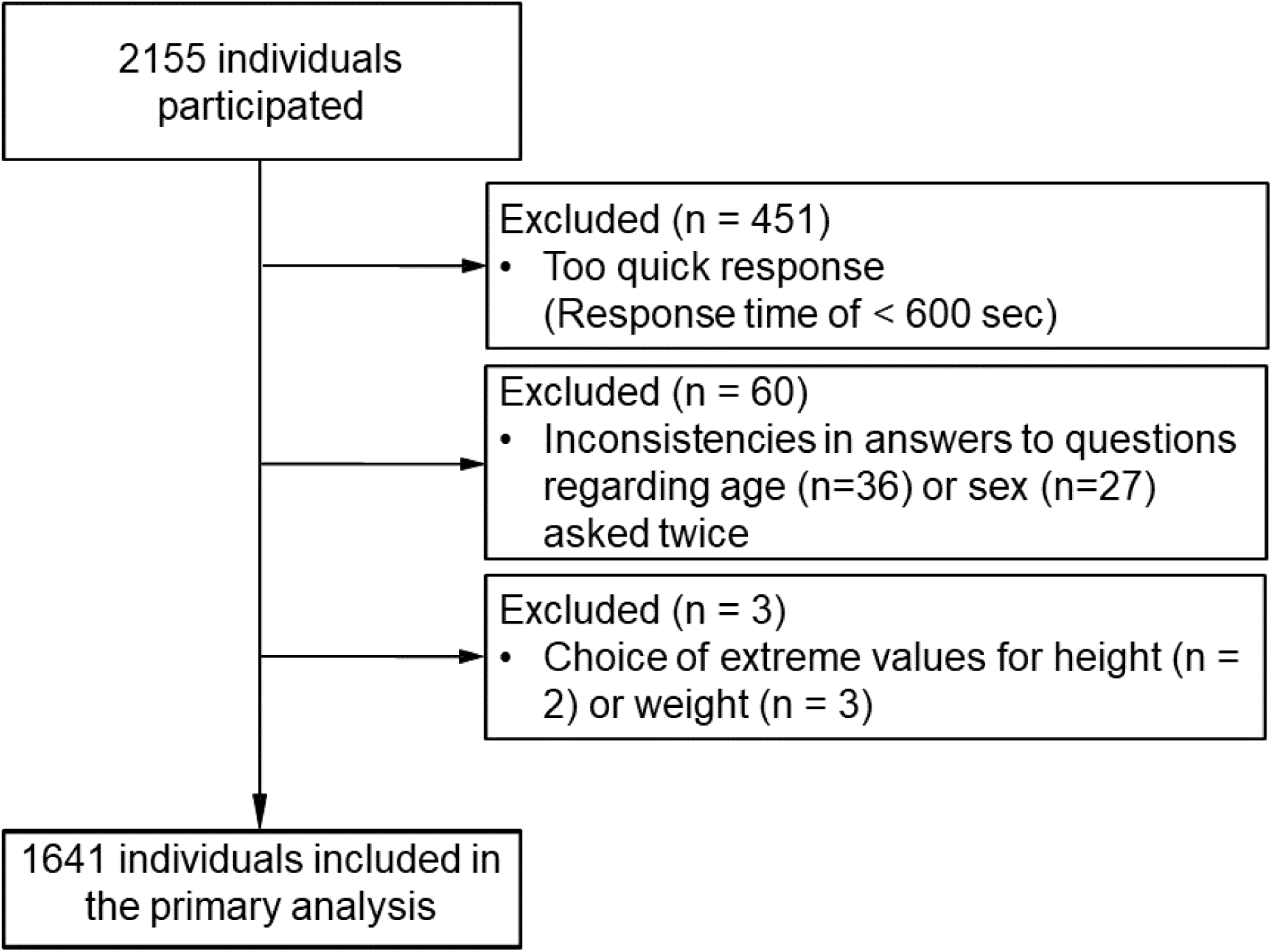
Flow of the study.

### Participant Characteristics

The mean age was 60.6 years, and 855 (52.1%) were women (Table 1). The reported psychiatric conditions included depression (12.5%), eating disorders (2.4%), and other mental illnesses (9.5%). The median BMI was 22.2 kg/m² (5th–95th percentile: 19.8–25.3), with 13.0% categorized as underweight, 60.2% as normal weight, 18.7% as overweight, and 8.0% as obese.

**Table 1.** Participant characteristics by body image distortion (N = 1641)

|  | Body image distortion |  |  | Total |
| --- | --- | --- | --- | --- |
|  | *No distortion* | *Underestimate* | *Overestimate* |  |
|  | n = 880 | n = 159 | n = 602 | N = 1641 |
| Demographics |  |  |  |  |
| Age, yr | 60.5 (12.8) | 58.7 (13.1) | 61.2 (12.9) | 60.6 (12.9) |
| Women, n (%) | 434 (49.3 %) | 51 (32.1 %) | 370 (61.5 %) | 855 (52.1 %) |
| BMI, kg/m<sup>2</sup> | 21.8 [19.5,26.4] | 21 [19.3,30.7] | 22.5 [20.8,23.9] | 22.2 [19.8,25.3] |
| *Underweight* | 125 (14.2 %) | 0 (0.0 %) | 89 (14.8 %) | 214 (13.0 %) |
| *Normal* | 416 (47.3 %) | 101 (63.5 %) | 471 (78.2 %) | 988 (60.2 %) |
| *Overweight* | 261 (29.7 %) | 4 (2.5 %) | 42 (7.0 %) | 307 (18.7 %) |
| *Obesity* | 78 (8.9 %) | 54 (34.0 %) | 0 (0.0 %) | 132 (8.0 %) |
| Education, n (%) |  |  |  |  |
| *Junior high school* | 28 (3.2 %) | 4 (2.5 %) | 17 (2.8 %) | 49 (3.0 %) |
| *High school* | 277 (31.5 %) | 57 (35.9 %) | 205 (34.1 %) | 539 (32.9 %) |
| *Professional training college/College of technology/Junior college* | 186 (21.1 %) | 28 (17.6 %) | 138 (22.9 %) | 352 (21.5 %) |
| *University* | 356 (40.5 %) | 58 (36.5 %) | 227 (37.7 %) | 641 (39.1 %) |
| *Graduate school* | 33 (3.8 %) | 12 (7.6 %) | 15 (2.5 %) | 60 (3.7 %) |
| Household income, n (%) |  |  |  |  |
| *<1,000,000 yen* | 80 (9.1 %) | 14 (8.8 %) | 44 (7.3 %) | 138 (8.4 %) |
| *1,000,000-<5,000,000 yen* | 489 (55.6 %) | 94 (59.1 %) | 319 (53.0 %) | 902 (55.0 %) |
| *5,000,000-<10,000,000 yen* | 229 (26.0 %) | 41 (25.8 %) | 179 (29.7 %) | 449 (27.4 %) |
| *>10,000,000 yen* | 82 (9.3 %) | 10 (6.3 %) | 60 (10.0 %) | 152 (9.3 %) |
| Marital status, n (%) |  |  |  |  |
| *Unmarried* | 187 (21.3 %) | 39 (24.5 %) | 111 (18.4 %) | 337 (20.5 %) |
| *Married* | 562 (63.9 %) | 95 (59.8 %) | 392 (65.1 %) | 1049 (63.9 %) |
| *Divorced* | 89 (10.1 %) | 18 (11.3 %) | 57 (9.5 %) | 164 (10.0 %) |
| *Widowed* | 42 (4.8 %) | 7 (4.4 %) | 42 (7.0 %) | 91 (5.6 %) |
| Regular exercise, n (%) | 281 (31.9 %) | 46 (28.9 %) | 194 (32.2 %) | 521 (31.8 %) |
| <u>Comorbidities</u> |  |  |  |  |
| Eating disorder, n (%) | 25 (2.8 %) | 4 (2.5 %) | 11 (1.8 %) | 40 (2.4 %) |
| Depression, n (%) | 122 (13.9 %) | 27 (17.0 %) | 56 (9.3 %) | 205 (12.5 %) |
| Other mental disorder, n (%) | 90 (10.2 %) | 27 (17.0 %) | 39 (6.5 %) | 156 (9.5 %) |
| <u>Cognitive-behavioral characteristics</u> |  |  |  |  |
| HLS-14 |  |  |  |  |
| Functional HL, pt | 18 [15, 21] | 18 [15, 22] | 18 [15, 21] | 18 [15, 21] |
| Communicative HL, pt | 18 [16, 20] | 18 [16, 20] | 18 [16, 20] | 18 [16, 20] |
| Critical HL, pt | 14 [12, 16] | 14 [12, 16] | 14 [12, 16] | 14 [12, 16] |
| TFEQ-R18V2 |  |  |  |  |
| Uncontrolled eating, pt | 29.6 [14.8,40.7] | 25.9 [11.1,37] | 29.6 [18.5,44.4] | 29.6 [14.8,40.7] |
| Emotional eating, pt | 16.7 [0,33.3] | 16.7 [0,33.3] | 22.2 [0,38.9] | 16.7 [0,33.3] |
| Cognitive restraint, pt | 33.3 [22.2,55.6] | 33.3 [0,55.6] | 44.4 [22.2,55.6] | 33.3 [22.2,55.6] |

<sup>a</sup>Means (SD) or medians [IQR] are presented for continuous data

### Eating Behavior Scale (TFEQ-R18V2): Descriptive and Psychometric Properties

The three-factor structure of the TFEQ-R18V2 demonstrated an acceptable fit for the data (RMSEA = 0.073, CFI = 0.934; see Additional File 6). Internal consistency was satisfactory across the domains, with Cronbach’s alpha values of 0.79 for CR, 0.89 for UE, and 0.92 for EE. The lowest standardized factor loading was 0.405 for Item 18, which was deemed acceptable given the overall model fit and reliability (Additional File 7).

Domain scores (mean ± SD) were 37.4 ± 23.7 for CR, 30.1 ± 19.3 for UE, and 22.4 ± 22.2 for EE. Although three EE items showed more than 50% responses at the lowest end of the scale, no notable floor effects were identified among participants who were overweight or obese (Additional File 8).

The expected correlation patterns with the DEBQ supported construct validity. Specifically, UE correlated moderately with DEBQ emotional and external eating, and weakly with restrained eating. CR showed a strong correlation with DEBQ-restrained eating and a weaker correlation with the other domains. EE was strongly correlated with emotional eating, moderately correlated with external eating, and weakly correlated with restrained eating (Additional File 9). All three TFEQ-R18V2 domains showed weak positive correlations with BMI. Collectively, these findings support this scale’s construct validity.

### Association between HL and BID

The median (interquartile range [IQR]) scores of health literacy (HL) were 18 [15, 21] for functional HL, 18 [16, 20] for communicative HL, and 14 [12, 16] for critical HL (Table 1).

With regard to BID, 602 participants (36.7%) overestimated their body size, 880 (53.6%) showed no distortion, and 159 (9.7%) underestimated their body size. Figure 2 illustrates the distribution of the body weight categories in the BID group. Among those who overestimated, approximately 80% were of normal weight, followed by 14.8% who were underweight. By contrast, approximately one-third of those who underestimated their body size were classified as obese.

**Figure 2.**
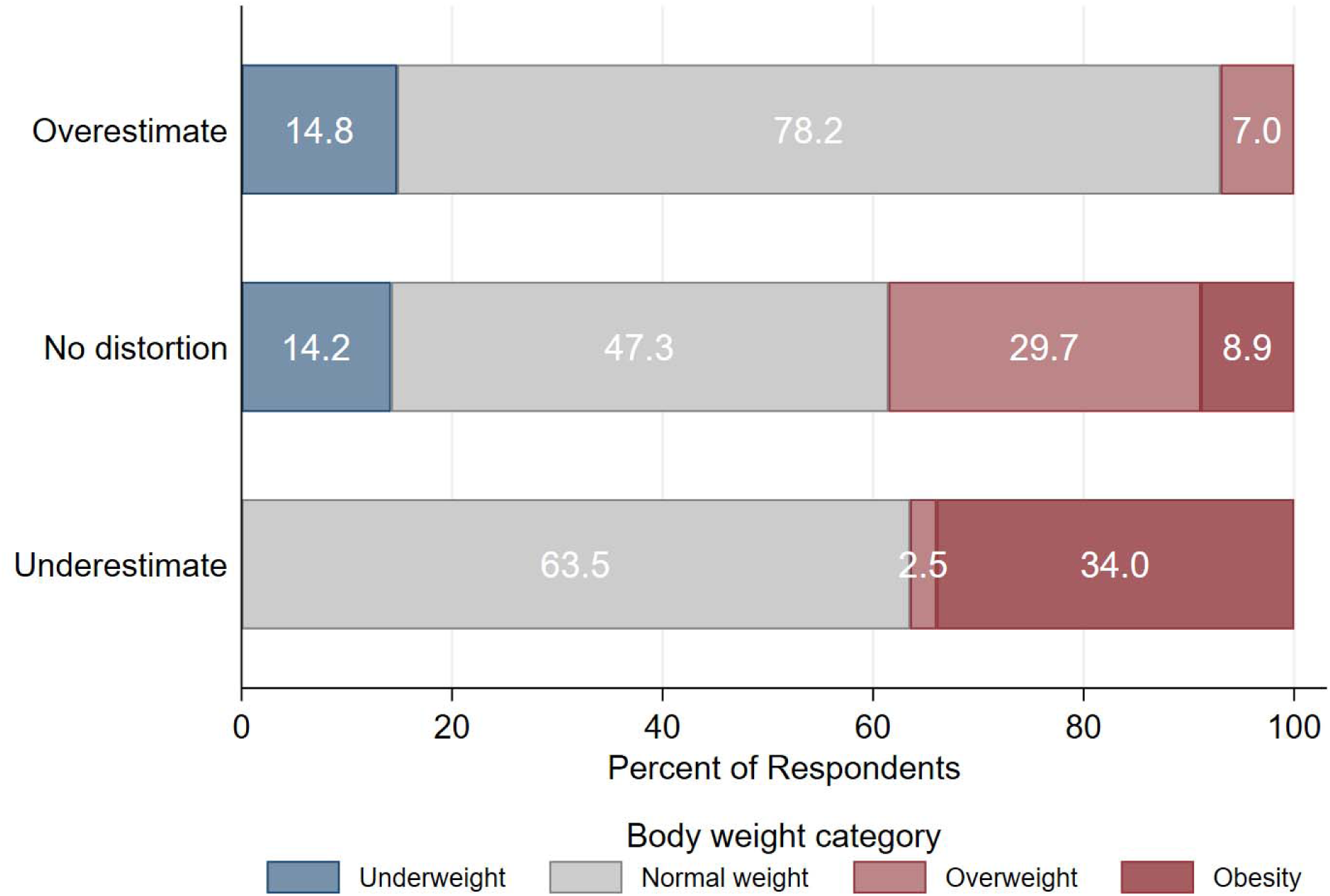
Proportion of Weight Categories by Type of Body Image Distortion.

Table 2 shows the associations among BID, HL scores, and patient characteristics. HL scores, whether functional, communicative, or critical, were not significantly associated with overestimation or underestimation.

**Table 2.** Associations Between Body Image Distortion and Health Literacy and Covariates (N = 1,641)

|  | Body image distortion (reference: no distortion) |  |  |  |
| --- | --- | --- | --- | --- |
|  | Overestimate<br>OR (95%CI) | P-value | Underestimate<br>OR (95%CI) | P-value |
| HLS-14, per 1-pt higher |  |  |  |  |
| Functional HL | 0.988 (0.963 - 1.014) | 0.380 | 1.008 (0.967 - 1.051) | 0.713 |
| Communicative HL | 0.984 (0.945 - 1.025) | 0.439 | 1.023 (0.957 - 1.094) | 0.501 |
| Critical HL | 1.005 (0.952 - 1.061) | 0.855 | 0.975 (0.895 - 1.062) | 0.559 |
| Age, per 10-yr higher | 1.017 (0.921 - 1.123) | 0.743 | 0.879 (0.747 - 1.035) | 0.121 |
| Women | **1.673 (1.320 - 2.119)** | **<0.001** | **0.415 (0.277 - 0.622)** | **<0.001** |
| Body mass index, per 1-kg/m<sup>2</sup> higher | **0.959 (0.935 - 0.983)** | **0.001** | 1.029 (0.992 - 1.066) | 0.125 |
| Education |  |  |  |  |
| Junior high school | Reference |  | Reference |  |
| High school | 1.070 (0.557 - 2.055) | 0.840 | 1.627 (0.536 - 4.937) | 0.390 |
| Professional training college/College of technology/Junior college | 0.979 (0.499 - 1.920) | 0.951 | 1.383 (0.437 - 4.381) | 0.581 |
| University | 1.015 (0.523 - 1.969) | 0.965 | 1.118 (0.362 - 3.446) | 0.847 |
| Graduate school | 0.817 (0.331 - 2.017) | 0.661 | 2.245 (0.614 - 8.208) | 0.221 |
| Household income |  |  |  |  |
| <1,000,000 yen | Reference |  | Reference |  |
| 1,000,000-<5,000,000 yen | 1.149 (0.763 - 1.732) | 0.506 | 1.178 (0.627 - 2.216) | 0.611 |
| 5,000,000-<10,000,000 yen | 1.501 (0.962 - 2.341) | 0.073 | 0.951 (0.472 - 1.917) | 0.889 |
| >10,000,000 yen | 1.470 (0.861 - 2.508) | 0.158 | 0.632 (0.252 - 1.587) | 0.329 |
| Marital status |  |  |  |  |
| Unmarried | Reference |  | Reference |  |
| Married | 1.002 (0.734 - 1.368) | 0.991 | 1.082 (0.669 - 1.749) | 0.749 |
| Divorced | 1.019 (0.662 - 1.571) | 0.930 | 1.212 (0.633 - 2.321) | 0.562 |
| Widowed | 1.284 (0.748 - 2.203) | 0.364 | 1.493 (0.578 - 3.857) | 0.408 |
| Regular exercise, yes | 0.961 (0.762 - 1.211) | 0.734 | 0.982 (0.666 - 1.447) | 0.926 |
| Eating disorder | 0.767 (0.347 - 1.694) | 0.511 | 0.756 (0.235 - 2.438) | 0.640 |
| Depression | 0.824 (0.547 - 1.242) | 0.356 | 0.916 (0.512 - 1.640) | 0.768 |
| Other mental disorder | 0.812 (0.507 - 1.301) | 0.386 | 1.674 (0.924 - 3.031) | 0.089 |

A multinomial logistic regression model was fitted, including all variables listed above. OR: odds ratio with “no distortion”
as the reference category; 95% CI: 95% confidence interval; HL: health literacy; HLS: Health Literacy Scale.

In contrast, women were positively associated with overestimation (adjusted odds ratio [aOR] 1.673; 95% confidence interval [CI], 1.320–2.119) and negatively associated with underestimation (aOR 0.415; 95% CI, 0.277–0.622). A higher BMI was inversely associated with overestimation (aOR per 1 kg/m² increase, 0.959; 95% CI, 0.935–0.983).

### Associations of Eating Behavior with HL and BID

Table 3 presents the associations among eating behavior, HL, and BID. Higher functional HL was associated with lower EE, UE, and CR. These associations remained consistent across the BID subgroups as no significant interactions were observed (P for interaction by the Wald test: 0.423, 0.989, and 0.758 for EE, UE, and CR, respectively).

**Table 3.**
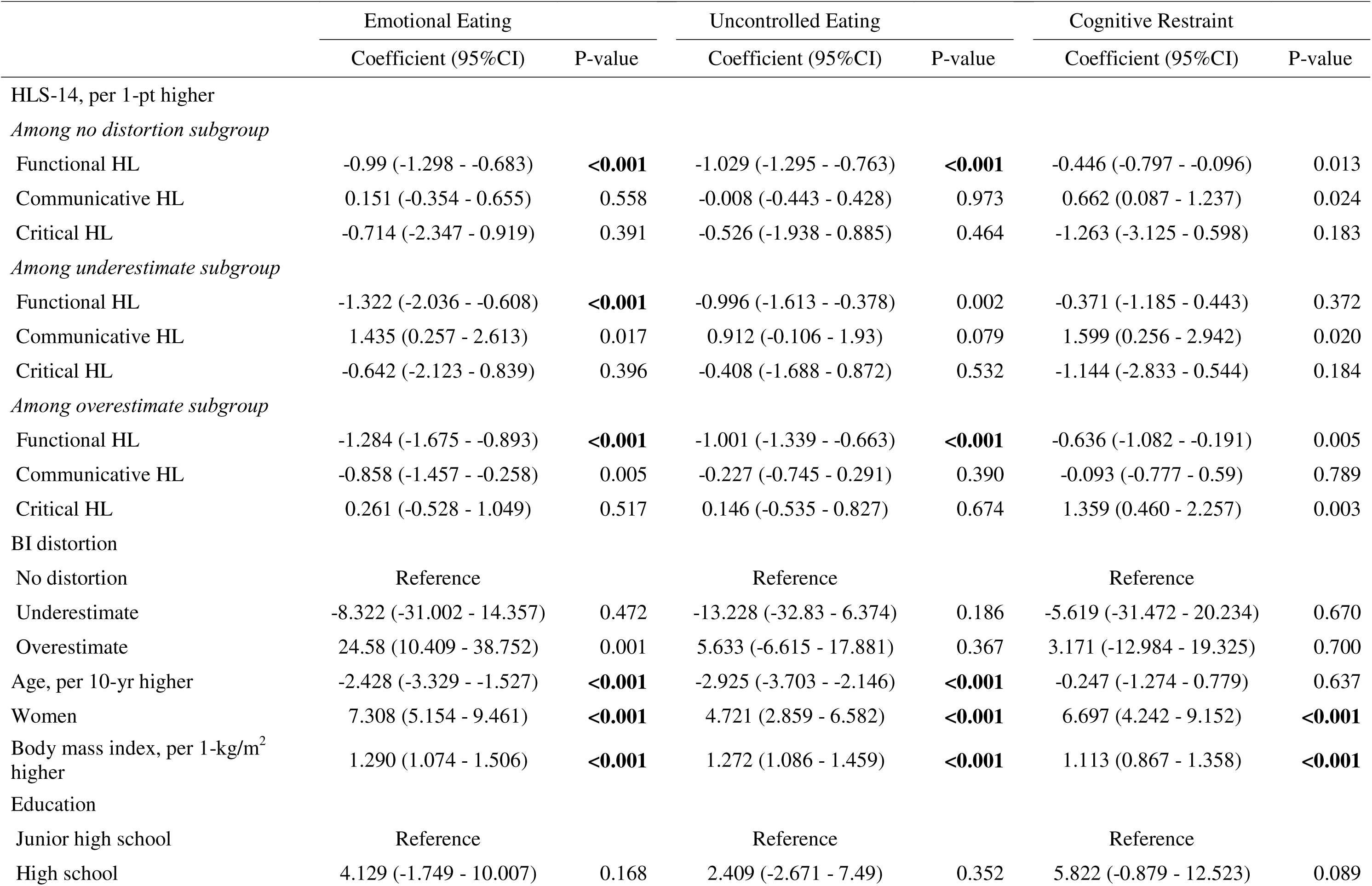
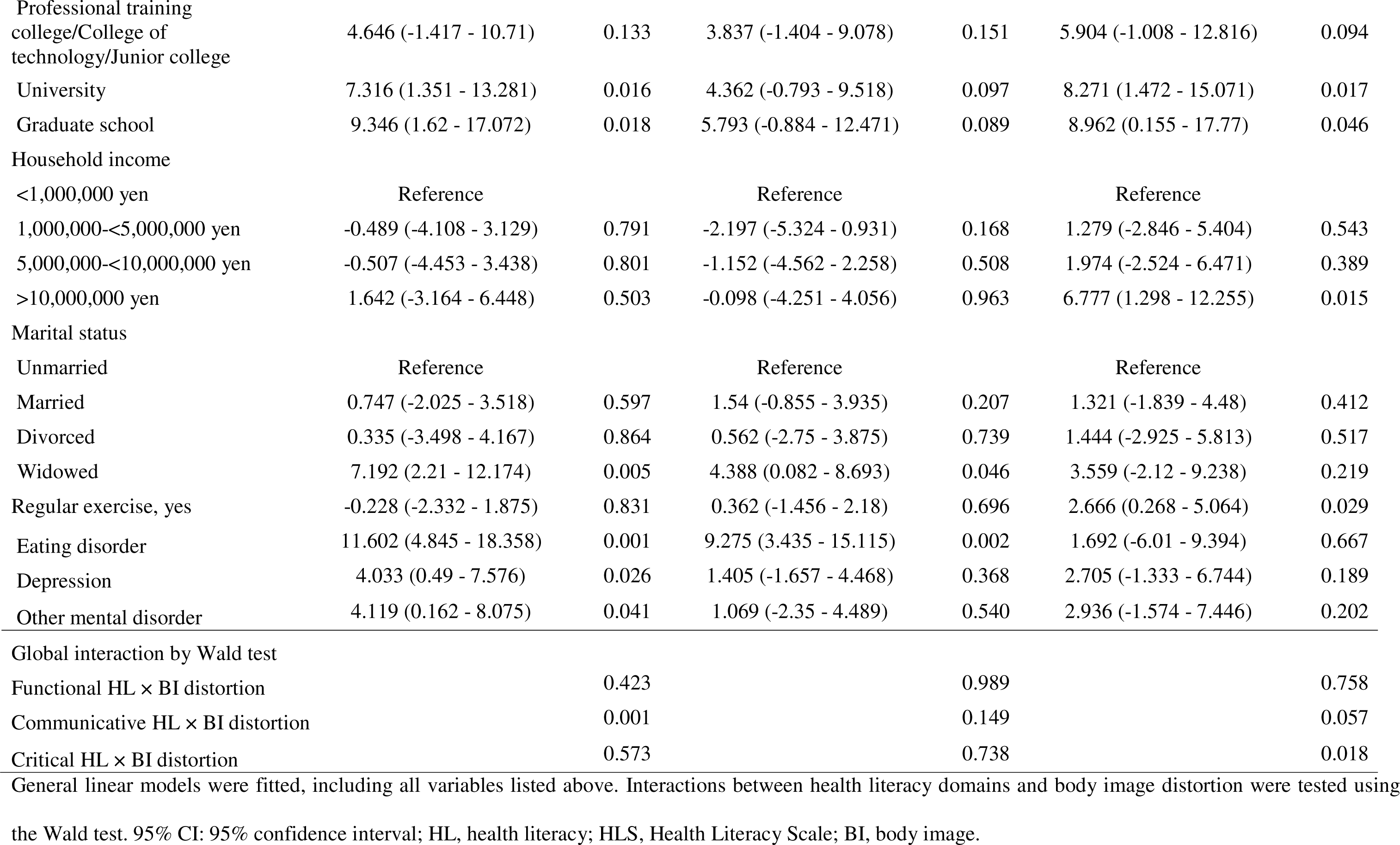
Associations Between Eating Behaviors and Health Literacy, Body Image Distortion, and Covariates (N = 1,641)

In contrast, the associations of communicative HL with both EE and CR were modified by BID (P for interaction: 0.001 for EE and 0.057 for CR). In the underestimate subgroup, higher communicative HL was positively associated with EE (per 1-point increase: β = 1.435; 95% CI: 0.257–2.613), whereas in the overestimate subgroup, it was negatively associated (β = –0.858; 95% CI: –1.457 to –0.258). Regarding CR, communicative HL showed a strong positive association in the underestimate subgroup (β = 1.599; 95% CI: 0.256–2.942) and a modest positive association in the no distortion subgroup (β = 0.662; 95% CI: 0.087–1.237).

Critical HL was also associated with CR in a BID-dependent manner (P for interaction = 0.018). Specifically, in the overestimate subgroup, higher critical HL was positively associated with CR (β = 1.359; 95% CI: 0.460–2.257).

Participants in the overestimate group showed significantly higher EE compared to those with no distortion (β = 24.58; 95% CI: 10.409–38.752). Lastly, higher BMI was positively associated with all three eating behavior domains: EE (β = 1.290; 95% CI: 1.074–1.506), UE (β = 1.272; 95% CI: 1.086–1.459), and CR (β = 1.113; 95% CI: 0.867–1.358).

## Discussion

Our findings suggest that enhancing functional HL may support healthier eating behaviors across subgroups with BID. Notably, communicative HL was associated with healthier eating patterns among individuals with overestimation-type BID but showed potentially adverse associations among those with underestimation. In contrast, critical HL was associated with more restrictive eating, specifically among individuals with overestimation, possibly reflecting more deliberate and reflective food choices.

Our finding that higher HL status was not associated with BID may initially appear inconsistent with previous studies; however, we believe it offers a more nuanced understanding of this relationship. A previous study of young adults in Iran found a positive correlation between higher HL and greater dissatisfaction or concern regarding BI [15]. However, that study included very few underweight participants, with nearly 40% classified as overweight and a notable proportion classified as obese. The observed association may have been driven by individuals with higher BMI expressing more concern about their BI; however, the analysis did not adjust for BMI. In contrast, our study controlled for BMI and found that a higher BMI was associated with less frequent overestimation of BI, suggesting that a lower BMI is linked to greater BI overestimation. Moreover, meta-analysis evidence has shown that interventions targeting media HL can reduce BI concerns among adolescents [27]. However, these studies primarily included Western adolescents aged 12–13 years, who may be more vulnerable to unreliable online health information owing to their developmental stage and limited health education. In this context, improving media HL may offer protection against BID. In contrast, the participants in our study were Japanese adults who had completed compulsory education by the age of 15 and likely received some degree of health education. Therefore, higher-order HL may not be associated with BID independent of sex or BMI in this population.

Consistent with previous studies, higher functional HL was associated with lower levels of undesirable eating behaviors, such as emotional and uncontrolled eating, regardless of the BID status. Similarly, research among Turkish adults has shown that adequate HL supports healthier food choices and overall eating behaviors [8]. Even among low-income populations, individuals with higher basic HL were more likely to avoid easily accessible high-fat foods such as fried chicken [28]. The observed association between higher functional HL and lower CR may reflect its buffering role against CR as an adaptive response to overweight or obesity [4]. This interpretation is supported by our finding that higher CR was positively associated with higher BMI.

A key contribution here is the novel insight that the association between higher-order HL, specifically communicative, and critical HL, and eating behavior, varies depending on the subtype of BID. This finding underscores the importance of tailoring HL interventions to individuals’ perception patterns. Notably, higher communicative HL was associated with lower EE only among individuals with an overestimated BID. This finding may reflect an increased awareness driven by aesthetic desires to lose weight, leading individuals to actively seek health information about food and regulate their eating behavior to avoid using food as a coping mechanism for negative emotions. Consequently, as seen in our sample, most individuals with an overestimation-type BID maintained their normal weight.

Interestingly, overestimation was strongly associated with higher EE, suggesting that those with unrealistic weight ideals may experience a gap between their actual and desired BI, which could lead to lower self-esteem and EE. This is consistent with previous findings that body dissatisfaction may increase EE, potentially through reduced self-esteem [29, 30]. Taken together, these results suggest that fostering communicative HL could be particularly beneficial for individuals with an overestimation-type BID in promoting healthier eating behaviors.

Conversely, among individuals with underestimation-type BID, higher communicative HL was paradoxically associated with higher EE. Given that one-third of this group was classified as obese, it is possible that while actively seeking health information, either from others or online, they encountered conflicting cues. For example, they may be confronted with external feedback that challenges their self-perception, or may be exposed to idealized body images that promote unrealistic standards and perfectionistic health behaviors. These experiences could contribute to self-criticism and, in turn, elevate EE. Another finding within the same subgroup supports this interpretation: higher communicative HL was also strongly associated with higher CR, suggesting an adaptive response. Notably, the positive association between communicative HL and CR was stronger among individuals with underestimation-type BID than among those without, further reinforcing the potentially adaptive nature of this behavior.

Interestingly, higher critical HL was associated with higher CR only among individuals with an overestimated BID. This suggests that excessive skepticism toward general health messages, such as the idea that moderation is key to healthy eating, may lead to overly restrictive behaviors. For example, driven by overestimation-type BID, young female dancers maintaining a healthy BMI have been reported to adopt restrictive eating patterns, such as vegan- or lactose-free diets [31]. Similarly, among Palestinians in their early 20s, those with higher nutrition-related critical HL were less likely to engage in behaviors such as adding cheese or mayonnaise to meals or spreading butter on bread [32].

The results here have several practical implications. First, regardless of the BID subtype, developing strategies to enhance functional HL may be beneficial. For individuals with limited HL, web-based nutrition education supervised by registered dietitians and focused on topics such as food groups and serving sizes may be more effective than printed materials [33]. Public health authorities can disseminate such resources using a one-way approach. Additionally, interactive educational workshops led by dietitians have been shown to improve food literacy, EE, and self-regulation, particularly among adolescents [34]. They may hold promise for broader application in the general adult population. Second, for individuals with an overestimation-type BID, interventions to enhance communicative HL, such as teaching-back techniques conducted by public health dietitians or psychologists, may help reduce EE [35]. By verifying their understanding and tailoring information accordingly, individuals may gain confidence in making healthy food choices and regulating their emotions. Simultaneously, for those with highly critical HL, public health professionals should create an open environment where individuals feel comfortable voicing their doubts. Providing clear and respectful explanations of eating behaviors could help prevent excessive dietary restrictions stemming from excessive skepticism. Finally, for individuals with underestimation-type BID, caution is warranted when promoting communicative HL. Unwanted or forced teaching-back interactions can harm self-esteem and trigger emotional or maladaptive restraint behaviors.

This study has several strengths. First, our large and demographically diverse Japanese sample, which included adults of both sexes and a broad age range, enhanced the generalizability of our findings. Second, the use of the HLS-14 allowed us to examine the roles of communicative and critical HL in addition to functional HL in supporting healthy eating behaviors across distinct BID subgroups.

This study had some limitations. First, BMI was based on self-reported height and weight, which may have underestimated the actual BMI. However, previous studies have shown strong correlations between self-reported and measured BMI (Spearman’s ρ > 0.9) [36], supporting its validity in epidemiological research. Second, owing to the cross-sectional design, reverse causality between HL and eating behaviors cannot be ruled out. Third, the HL scale focused on understanding health information in the context of illness and did not capture media- or nutrition-specific literacy. Further studies are needed to assess whether media literacy is associated with BID among Japanese adults.

## Conclusion

In conclusion, our study found that functional HL was associated with healthier eating behaviors, regardless of BID. However, the association between communicative and critical HL and eating behaviors differed according to BID subtype, particularly between overestimation and underestimation. Further research is needed to determine whether tailoring the level and delivery of HL interventions to individual BID patterns can promote healthy eating behaviors more effectively.

## Data Availability

The datasets generated and analyzed in the current study are available from the corresponding author upon reasonable request.

## List of Abbreviations

aOR: Adjusted odds ratio
BI: Body image
BID: Body image distortion
BMI: Body mass index
CFI: Comparative fit index
CR: Cognitive restraint
DEBQ: Dutch Eating Behavior Questionnaire
EE: Emotional eating
HLS-14: the 14-item health literacy scale
IQR: Interquartile range
RMSEA: Root mean square approximation error
SDs: Standard deviations
TFEQ: Three-Factor Eating Questionnaire
TFEQ-R18V2: Japanese version of the 18-item Three-Factor Eating Questionnaire-R18 version 2
UE: Uncontrolled eating

## Declarations

### Ethics approval and consent to participate

This cross-sectional online survey was approved by the Institutional Review Board at Fukushima Medical University (ippan2022-210). Only the participants who provided informed consent completed the questionnaire.

### Consent for publication

Not applicable

### Authors’ contributions

**NK** conceptualized the study, designed the methodology, analyzed the data, performed the statistical analyses, and wrote the original and final drafts. **TM** conceptualized the study, designed the methodology, performed the statistical analyses, and wrote the original draft. **SS** analyzed data. **TW** designed the methodology of the study. **TA** conceptualized this study. **HK** designed the methodology and analyzed the data. All the authors have read and approved the final manuscript.

## Acknowledgments

Not applicable

