## Additional file 1 for "Communicative and critical health literacy and eating behaviors in Japanese adults: the modifying role of body image distortion": Additional_file_1 Sampling Screener R.docx

### A. Participant Sampling Strategy

We recruited Japanese adults aged ≥20 years through a web-based survey company (Cross Marketing, Shinjuku, Tokyo, Japan). Due to budget constraints, the target sample size was set at 1,500 participants. Stratified sampling was used to achieve a 1:1 ratio by sex (750 men and 750 women) and age group (≥65 vs. <65 years), resulting in 375 individuals in each subgroup.

To include individuals with and without obesity, we applied predefined panel categories used by the survey company. Specifically, we sampled individuals based on self-reported obesity status to achieve a 10:3:2 ratio: (1) those with a history of medical visits or hospitalization for obesity, (2) those concerned about obesity, and (3) those with neither. For each sex and age group, this corresponded to 250, 75, and 50 individuals, respectively.

**Additional File 1.**

**B. Screener Item Design**

To identify and exclude careless respondents [[1]](https://paperpile.com/c/yiLwwt/rEPUf), we implemented five exclusion criteria:

(1) inconsistent or implausible entries for age or sex,

(2) extreme values for height or weight, and

(3) survey completion time under 5 minutes [[2, 3]](https://paperpile.com/c/yiLwwt/LGyDR+wMKeM).

Participants were asked to report their age and sex twice—once at the beginning and again near the end of the questionnaire. Respondents with inconsistent answers were excluded.

Height and weight were reported once, toward the end of the survey. To exclude implausible anthropometric data, we removed responses with height ≥210 cm or ≤120 cm, and weight ≥151 kg or ≤25 kg.

Based on a pilot test, we excluded respondents who completed the survey in under five minutes, as such rapid completion was considered indicative of careless responding [[1, 2]](https://paperpile.com/c/yiLwwt/rEPUf+LGyDR).
