## Additional file 2 for "Communicative and critical health literacy and eating behaviors in Japanese adults: the modifying role of body image distortion": Additional_file_2 FRS.docx

**Additional File 2. Japanese version of the Figure Rating Scale**

Participants were presented with the instruction: “Which figure do you think resembles yours? (CURRENT)” (Japanese: 「現在のあなたの容姿に似ていると思う図は何番ですか。」) [[1,2]](https://paperpile.com/c/yiLwwt/SLE7u+qloRS). Depending on their reported sex, participants were then shown either the female or male silhouette set and asked to select the number corresponding to the figure they felt most closely resembled their current body shape.

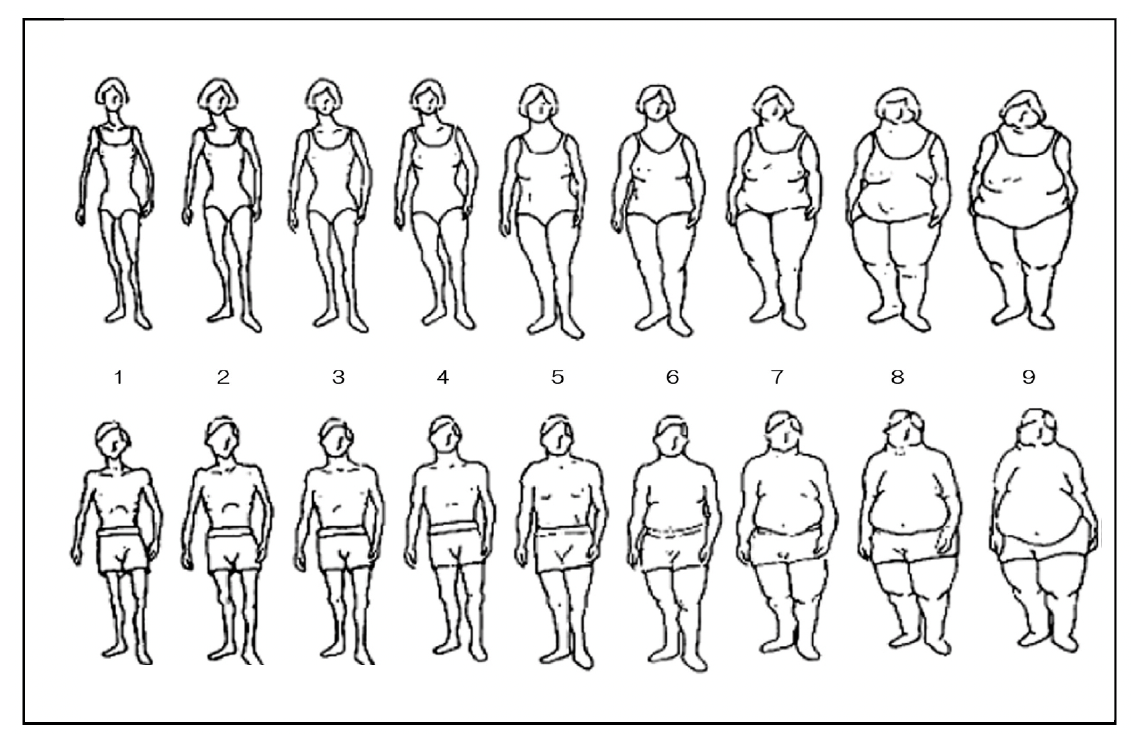
