## Additional file 3 for "Communicative and critical health literacy and eating behaviors in Japanese adults: the modifying role of body image distortion": Additional_file_3 TFEQ18Rv2 Refæ}ôné╠é╜é▀æ╥

**Additional File 3. Japanese version of The Three-Factor Eating Questionnaire-Revised 18-Item version 2 (TFEQ-R18V2)**

Questionnaires in Japanese version are available from the following website (<https://noriaki-kurita.jp/wp-content/uploads/2025/08/05579ae1a26b16c1f582442cb34ef870.pdf>)

| Instruction | *  Please circle the number for the response that best describes how much you agree or disagree with each of the following statements. |
| --- | --- |
| Question 1 | I deliberately take small helpings to control my weight. † |
| Question 2 | I start to eat when I feel anxious. † |
| Question 3 | Sometimes when I start eating, I just can’t seem to stop. † |
| Question 4 | When I feel sad, I often eat too much. † |
| Question 5 | I don’t eat some foods because they make me fat. † |
| Question 6 | Being with someone who is eating often makes me want to also eat. † |
| Question 7 | When I feel tense or ‘‘wound up’’, I often feel I need to eat. † |
| Question 8 | I often get so hungry that my stomach feels like a bottomless pit. † |
| Question 9 | My doctor is well qualified to manage (diagnose and treat or make an appropriate referral) medical problems like mine. † |
| Question 10 | When I feel lonely, I console myself by eating. † |
| Question 11 | I consciously hold back on how much I eat at meals to keep from gaining weight. † |
| Question 12 | When I smell a sizzling steak or see a juicy piece of meat, I find it very difficult to keep from eating - even if I've just finished a meal. † |
| Question 13 | I’m always hungry enough to eat at any time. † |
| Question 14 | If I feel nervous, I try to calm down by eating. † |
| Question 15 | When I see something that looks very delicious, I often get so hungry that I have to eat right away. † |
| Question 16 | When I feel depressed, I want to eat. † |
| Response options for Questions 1 to 16 | Definitely true/Mostly true/Mostly false/Definitely false |
| Question 17 | Do you go on eating binges even though you’re not hungry? |
| Response options for Question 17 | Never/Rarely/Sometimes/At least once a week |
| Question 18 | How often do you feel hungry? |
| Response options for Question 18 | Only at mealtimes/Sometimes between meals/Often between meals/Almost always |

The original English version [[1]](https://paperpile.com/c/yiLwwt/WYRSU) is also provided for each item and response.

*The instructional statements generated through the formal process of translation are presented.

†Questions 1 through Question 16 are reverse-scored items.

Using the eighteen questions, the following three domains can be assessed:

**Uncontrolled eating (UE)**: Question 3, Question 6, Question 8, Question 9, Question 12, Question 13, Question 15, Question 17, and Question 18.

**Cognitive restraint (CR)**: Question 1, Question 5, Question 11.

**Emotional eating (EE)**: Question 2, Question 4, Question 7, Question 10, Question 14, and Question 16

Before using this instrument, please register at https://noriaki-kurita.jp/resources/tfeq-r21-jpn/.

In addition, please cite this article as follows:

Kurita N, Maeshibu T, Shimizu S, Aita T, Wakita T, Kikuchi H. Communicative and critical health literacy and eating behaviors in Japanese adults: the modifying role of body image distortion. *medRxiv*. 2025:2025.XX.XX.YYYYYYYY. DOI
