## Additional file 4 for "Communicative and critical health literacy and eating behaviors in Japanese adults: the modifying role of body image distortion": Additional_file_4 Survey items.docx

**Additional File 4. Survey Items Used in the Present Study**

The Japanese version of the Dutch Eating Behavior Questionnaire (DEBQ) is a 33-item instrument assessing three eating behavior domains: emotional eating (13 items), referring to eating in response to emotional arousal; external eating (10 items), triggered by external cues such as taste or smell; and restrained eating (10 items), reflecting intentional dietary restriction [[1, 2]](https://paperpile.com/c/yiLwwt/qxkoe+eUMPI). Participants rated each item on a 5-point Likert scale (1 = never to 5 = very often), and mean scores were calculated for each domain. Higher scores indicated greater expression of the corresponding eating behavior. Internal consistency (Cronbach’s alpha) was 0.95 for emotional eating, 0.73 for external eating, and 0.87 for restrained eating [[2]](https://paperpile.com/c/yiLwwt/eUMPI). Criterion validity in Japanese adults showed a weak negative correlation between external eating and BMI, a weak positive correlation between restrained eating and BMI, and no significant correlation with emotional eating [[2]](https://paperpile.com/c/yiLwwt/eUMPI).

Covariates included demographic characteristics (age, sex, education level, household income, marital status), health behaviors (exercise, smoking, alcohol consumption), and history of psychiatric conditions (eating disorders, depression, other mental disorders).

Sex was reported as either male or female. The educational level was junior high school, high school, professional training college, technology college, junior college, university, or graduate school. For the analyses, the data were collapsed into five levels (junior high school, high school, professional training college/college of technology/junior college, university, and graduate school). Total household income was classified as < 1 000 000 yen; 1 000 000 to < 2 999 999 yen; 3 000 000 to < 4 999 999 yen; 5 000 000 to < 9 999 999 yen; ≥ and 10,000,000 yen. For the analyses, the data were collapsed into four levels < 1 000 000 yen; 1 000 000 to < 2 999 999 yen; 3 000 000 to < 4 999 999 yen; 5 000 000 to < 9 999 999 yen; ≥ and 10,000,000 yen). Marital status was recorded as unmarried, married, divorced, or widowed.

Health behaviors were assessed using items adapted from the Ministry of Health, Labour and Welfare’s standard questionnaire for specific health checkups in Japan [[3]](https://paperpile.com/c/yiLwwt/adzLH). Regular exercise was defined as answering “yes” to the question: “I engage in light, sweaty exercise for at least 30 minutes on two or more days a week and have continued this for at least one year.” This corresponds to ≥60 minutes of moderate activity (≥3 METs) per week, which is associated with a 12% reduction in the risk of lifestyle-related diseases and mortality [[3]](https://paperpile.com/c/yiLwwt/adzLH).

Psychiatric history was assessed by asking: “Have you ever been told by a physician that you have any of the following conditions?” (eating disorders, depression, or other mental disorders). Response options were: “never told,” “told in the past,” or “told and currently receiving care.” Respondents selecting either of the latter two options were considered to have the condition.
