## Additional file 5 for "Communicative and critical health literacy and eating behaviors in Japanese adults: the modifying role of body image distortion": Additional_file_5 Stat descript Spearman corrected.docx

**Additional File 5. Detailed description of statistical analyses**

Psychometric analyses were performed using R version 4.1.2, the psych package version 2.2.3, and the lavaan package version 0.6-11. All other analyses were performed using Stata/SE. version 17 (Stata Corp., College Station, TX, USA). Respondent characteristics are presented as means and standard deviations or medians with interquartile ranges for continuous variables, and as frequencies and percentages for categorical variables.

For the TFEQ-R18V2, three-factor confirmatory factor analyses were conducted to examine the goodness of fit of the three-factor model. Recorded raw scores were used (items 1–16). The goodness of fit of the models was assessed using the comparative fit index (CFI) and root mean square approximation error (RMSEA). Acceptable model criteria are a CFI ≥ 0.90 and an RMSEA ≤ 0.08 [[1]](https://paperpile.com/c/yiLwwt/YpRis). Acceptable standardized loadings were set at ≥ 0.3 while also considering the goodness of fit of the model and internal consistency reliability [[2]](https://paperpile.com/c/yiLwwt/5Qc8A). The distributions of the responses were examined at the item level. Positive floor (>50% of the responses at the lower end of the possible scores) and ceiling effects (>50% of the responses at the upper end of the possible scores) were examined [[3]](https://paperpile.com/c/yiLwwt/WYRSU). The reliability of each domain in the three-factor models was assessed by Cronbach's α and McDonald's ω coefficients [[4]](https://paperpile.com/c/yiLwwt/CYRR7). In general, coefficients > 0.7 are recommended [[4]](https://paperpile.com/c/yiLwwt/CYRR7).

Furthermore, construct validity was examined by testing the correlations between the TFEQ domain scores and the Japanese version of the DEBQ and BMI. Spearman correlation coefficient was calculated to examine correlations.

We hypothesized that the UE domain of the TFEQ-R18V2, which reflects general difficulties in regulating eating, including hunger, would show at least moderate correlations with the “External Eating” and “Emotional Eating” domains of the DEBQ. In contrast, its correlation with the “Restrained Eating” domain was expected to be weak or negligible. The CR domain, which represents conscious restriction of food intake to control weight, was expected to be strongly correlated with the DEBQ “Restrained Eating” domain and weakly or negligibly correlated with the “External Eating” and “Emotional Eating” domains. The EE domain, which captures overeating in response to negative emotions, was expected to be strongly correlated with the DEBQ “Emotional Eating” domain and weakly or negligibly correlated with the “External Eating” and “Restrained Eating” domains.

In a previous web-based survey conducted in the original validation study, all three TFEQ domains (UE, CR, and EE) showed weak positive correlations with BMI [[3]](https://paperpile.com/c/yiLwwt/WYRSU). Based on this, we expected similar patterns in the present study. The weak positive correlation between CR and BMI may suggest that excessive dietary restraint could paradoxically lead to episodes of overeating.
