## Additional file 6 for "Communicative and critical health literacy and eating behaviors in Japanese adults: the modifying role of body image distortion": Additional_file_6 structural validity 18RV2.docx

**Additional File 6. Three-factor structural validity and internal consistency reliability of the domains in the TFEQ-R18V2**

| **Goodness-of-fit indices based on confirmatory factor analysis** |  |
| --- | --- |
| Comparative Fit Index | 0.934 |
| Root Mean Square Error of Approximation | 0.073 |
| Standardized Root Mean Square Residual | 0.050 |
| **Internal consistency reliability** |  |
| Cronbach's α coefficient |  |
| Uncontrolled Eating | 0.89 |
| Cognitive Restraint | 0.79 |
| Emotional Eating | 0.92 |
| McDonald's ω coefficient |  |
| Uncontrolled Eating | 0.89 |
| Cognitive Restraint | 0.80 |
| Emotional Eating | 0.93 |
