## Additional file 7 for "Communicative and critical health literacy and eating behaviors in Japanese adults: the modifying role of body image distortion": Additional_file_7 CFA of 18RV2.docx

### Additional File 7. Confirmatory factor analysis for TFEQ-R18V2

Dark grey squares indicate the items. Dark grey circles indicate domains. Digits overlaid on the one-way arrows indicate standardized loadings. The digits overlaid on the two-way arrows indicate the correlations.

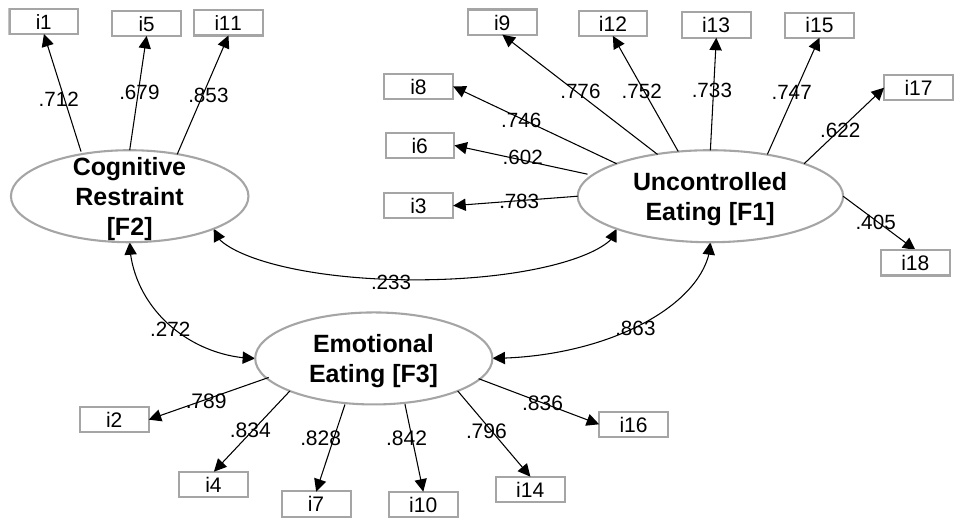
