## Additional file 8 for "Communicative and critical health literacy and eating behaviors in Japanese adults: the modifying role of body image distortion": Additional_file_8 Floor and ceiling 18RV2.docx

### Additional File 8. Flooring and ceiling by item

| **Total population (N = 1641)** | | | | |  | **Overweight/Obesity subgroup (n = 439)** | | | | |
| --- | --- | --- | --- | --- | --- | --- | --- | --- | --- | --- |
| % of flooring | |  | % of ceiling | |  | % of flooring | |  | % of ceiling | |
| Item | % |  | Item | % |  | Item | % |  | Item | % |
| 14R | 55.2 % |  | 6R | 7.3 % |  | 14R | 41.2 % |  | 6R | 12.1 % |
| 4R | 53.4 % |  | 11R | 5.9 % |  | 4R | 39.2 % |  | 3R | 10.5 % |
| 10R | 52.3 % |  | 3R | 5 % |  | 10R | 38.5 % |  | 8R | 7.7 % |
| 16R | 46.8 % |  | 5R | 4.9 % |  | 16R | 33.7 % |  | 7R | 7.1 % |
| 2R | 45.3 % |  | 8R | 4.3 % |  | 13R | 31.2 % |  | 2R | 6.6 % |
| 13R | 45.2 % |  | 7R | 4 % |  | 2R | 29.8 % |  | 17 | 5.9 % |
| 7R | 45 % |  | 2R | 3.8 % |  | 7R | 29.6 % |  | 5R | 5.7 % |
| 9R | 43.3 % |  | 1R | 3.3 % |  | 5R | 28 % |  | 11R | 5.5 % |
| 8R | 41.3 % |  | 4R | 3 % |  | 18 | 27.1 % |  | 4R | 5 % |
| 3R | 37.2 % |  | 17 | 3 % |  | 9R | 26.4 % |  | 15R | 4.8 % |
| 5R | 36.8 % |  | 15R | 2.8 % |  | 8R | 24.6 % |  | 16R | 4.3 % |
| 17 | 33.8 % |  | 16R | 2.6 % |  | 15R | 19.8 % |  | 10R | 4.1 % |
| 15R | 33.3 % |  | 12R | 2.6 % |  | 12R | 19.6 % |  | 12R | 4.1 % |
| 18 | 32.9 % |  | 10R | 2.4 % |  | 3R | 18.7 % |  | 9R | 3.6 % |
| 12R | 32.2 % |  | 18 | 2.3 % |  | 17 | 17.1 % |  | 18 | 3.6 % |
| 1R | 22.9 % |  | 9R | 2 % |  | 1R | 14.8 % |  | 1R | 2.7 % |
| 11R | 21.1 % |  | 14R | 1.7 % |  | 6R | 10 % |  | 14R | 2.5 % |
| 6R | 16.8 % |  | 13R | 1.7 % |  | 11R | 8.7 % |  | 13R | 2.3 % |

R indicates the re-coded score

Items 4, 10, and 14 are components of the emotional eating subdomain:

4. When I feel sad, I often eat too much.

10. When I feel lonely, I console myself by eating.

14. If I feel nervous, I try to calm down by eating.
