## Additional file 9 for "Communicative and critical health literacy and eating behaviors in Japanese adults: the modifying role of body image distortion": Additional_file_9 Criterion validity Spearman 18RV2.docx

**Additional File 9. Criterion validity of the TFEQ-R18V2**

|  | DEBQ | | |  |
| --- | --- | --- | --- | --- |
|  | External eating | Restrained eating | Emotional eating | BMI |
| TFEQ-R18V2 |  |  |  |  |
| Uncontrolled eating | 0.6327; p <0.001 | 0.1277; p <0.001 | 0.6735; p <0.001 | 0.3148; p <0.001 |
| Cognitive restraint | 0.1190; p <0.001 | 0.7231; p <0.001 | 0.2311; p <0.001 | 0.2557; p <0.001 |
| Emotional eating | 0.4825; p <0.001 | 0.1452; p <0.001 | 0.7828; p <0.001 | 0.2752; p <0.001 |

Spearman correlation coefficients between the three factors in the TFEQ-R21 and the three factors in the DBEQ and BMI are shown.

TFEQ-R18V2, The 18-item Three-Factor Eating Questionnaire, version 2; DEBQ, Dutch Eating Behavior Questionnaire; BMI, body mass index.
